# Moderate to vigorous physical activity and sedentary behavior change in self-isolating adults during the COVID-19 pandemic in Brazil: A cross-sectional survey exploring correlates

**DOI:** 10.1101/2020.07.15.20154559

**Authors:** Felipe B. Schuch, Rugero A. Bulzing, Jacob Meyer, Guillermo F. López-Sánchez, Igor Grabovac, Peter Willeit, Davy Vancampfort, Cristina M. Caperchione, Kabir P. Sadarangani, André O. Werneck, Philip B. Ward, Mark Tully, Lee Smith

## Abstract

**Background:** Self-distancing measures imposed major changes in daily life. This study aimed to (i) evaluate the changes (pre-versus during pandemic) in time spent in moderate to vigorous physical activity (MVPA) and sedentary behavior (SB) in self-isolating Brazilians during the COVID-19 pandemic, and (ii) to explore correlates of MVPA and SB.

**Methods:** A cross-sectional self-report online survey, evaluating the time spent in MVPA and SB pre and during the COVID-19 pandemic. Sociodemographic, behavioral, clinical, variables, and time in self-isolation were also obtained. Changes in MVPA and SB and their correlates were explored using generalized estimating equations (GEE).

**Results:** A total of 877 participants (72.7% women, 53.7% young adults [18-34 years]) were included. Overall, participants reported a 59.7% reduction (95%CI:35.6 to 82.2) in time spent on MVPA during the pandemic. Time spent in SB increased 42.0% (95%CI:31.7 to 52.5). Greater reductions in MVPA and/or increases in SB were seen in younger adults, those not married, those employed and those with a self-reported previous diagnosis of a mental disorder.

**Conclusions:** People in self-isolation significantly reduced MVPA levels and increased SB. Public health strategies should be implemented during epidemic times to mitigate the impact of self-isolation on MVPA and SB, particularly in vulnerable groups.

## Introduction

The coronavirus disease 2019 (COVID-19) pandemic, caused by the spread of the severe acute respiratory coronavirus 2 (SARS-CoV-2), has infected more than 13 million people in more than 200 countries around the world resulting in nearly 570 thousand deaths on the 14^th^ of July 2020 according to the World Health Organization (WHO)^1^. As a response to reduce the virus spread, the WHO recommended national governments to adopt non-pharmacological strategies based on social and physical distancing, such as lockdown, quarantine, and self-isolation.

In Brazil, the epidemiological report number 05 of the Ministry of Health has recommended the adoption of social distancing measures, including self-isolation in areas with community transmission^2^. When self-isolating, people were advised to stay at home, and only go out in public for essential activities, such as going to the supermarket, pharmacy, or to use essential services, such as medical assistance. All other non-essential services, including gyms, parks, stadiums, and other places where people exercised were closed.

Self-isolation measures impose a drastic and sudden disruption of daily life routine, resulting in limited physical and social mobility, and fewer opportunities to be active^3^. Moreover, the emotional burden^4, 5^ of the pandemic likely resulted in additional barriers to remaing focused and motivated to be and/or stay physically active, potentially reducing the time spent in physical activity (PA), defined as any bodily movement produced by skeletal muscles that result in energy expenditure^6^. The negative impact of the pandemic and self-isolation measures on PA levels, both on light PA and moderate to vigorous PA (MVPA), was noted in many countries including Australia^7, 8^, Canada^9^, Croatia^10^, France^11^, Italy^12^, Spain^13^, the UK^14, 15^, and the USA^16-18^. However, some moderating factors on PA changes in this period were identified. For example, age^12, 14^, sex^13^ and the presence of chronic physical diseases or mental disorders moderated the pandemic impact on PA levels^14^. Further, increases in time spent in sedentary behavior (SB), defined as any waking behavior characterized by an energy expenditure ≤1.5 metabolic equivalents (METs), while in a sitting, reclining or lying posture^19^, were noted during the pandemic in the US^17, 18^, France and Sweden^20^, and Spain^13^. In Spain, sex moderated the increase in time spent in SB^13^.

To the best of our knowledge, no study has evaluated how the COVID-19 pandemic changed PA and SB during self-isolating Brazilians. The present study aimed (i) to examine the changes (pre-versus during COVID-19 pandemic) on PA and SB in self-isolating Brazilians, and (ii) to evaluate whether sociodemographic (sex, age, ethnicity, marital status, employment, monthly household income), behavioral (smoking, current alcohol consumption), clinical (presence of chronic physical diseases or mental disorders), and contextual factors (i.e., number of days of self-isolation) moderated these changes.

## Methods

This paper presents pre-planned interim analysis of data from a cross-sectional study. Data collection was performed through an online survey (www.qualtrics.com). The study was launched on 11 April 2020 and data collection continued until 05 May. The study was approved by the Federal University of Santa Maria Research Ethics Committee and by the National Commission of Ethics in Research [CONEP] (30244620.1.0000.5346).

Participants were recruited through social media (Facebook, Instagram, Twitter), and by distributing an invitation to participate through existing researcher networks. The inclusion criteria were: (1) Brazilians adults (18-65 years), (2) currently residing in Brazil, and (3) in self-isolation due to the COVID-19 pandemic. Self-isolation was defined as staying-at-home and only leaving for essential activities such as food shopping, visiting the pharmacist or other health professionals. Participants who self-reported the presence of COVID-19 symptoms, assessed through a list of symptoms (fever, cough, dry mouth, coriza, sore throat), were removed from this analysis (n=65).

### Moderate to vigorous physical activity (MVPA) and sedentary behavior (SB) assessment

Participants were asked to recall the amount of time in vigorous and moderate physical activity, and sedentary behavior they undertook on an average day, separately both pre- and since self-isolation^15^. Participants were asked: (1) “How much time on an average day have you spent in vigorous activity before/since social distancing?”; (2) “How much time on an average day have you spent in moderate activity since/before social distancing?” and (3) “How much time on an average day have you spent sitting since/before social distancing?” Responses were given in hours and minutes. MVPA and SB were analyzed as continuous variables (minutes per day). We also categorized PA levels (≥30 minutes/day or <30 minutes/day of MVPA), which is in accordance with the WHO recommendations. Next, four categories were derived to identify patterns of change: (1) persistent inactive (< 30 minutes/day of MVPA pre and during the pandemic), (2) decreased PA (≥30 minutes/day of MVPA pre and <30 minutes/day of MVPA during the pandemic), (3) increased PA (<30 minutes/day of MVPA pre and ≥30 minutes/day of MVPA during the pandemic) and (4) persistent active (≥30 minutes/day of MVPA pre and during the pandemic).

### Covariates

Demographic data were collected, including sex (men or women), age (in 10- year age bands), ethnicity (Caucasian, Black, Asian, mixed, others), marital status (single, divorced, widowed or married), employment (employed, student, military, unemployed), monthly household income ≤R$1254, R$1255-R$2004, R$2005-R$8640, R$8641-R$11261 ≥R$11262). Health behaviors data included current smoking (y/n) or alcohol consumption (y/n). Clinical data included self-reported previous diagnosis of physical diseases or mental disorders, such as: obesity, hypertension, myocardial infarction, angina pectoris, and other coronary diseases, other cardiac diseases, varicose veins of lower extremities, osteoarthritis, chronic neck pain, chronic low back pain, chronic allergy (excluding allergic asthma), chronic bronchitis, emphysema or chronic obstructive pulmonary disease, type 1 diabetes, type 2 diabetes, diabetic retinopathy, cataract, peptic ulcer disease, urinary incontinence or urine control problems, hypercholesterolemia, chronic skin disease, chronic constipation, liver cirrhosis and other hepatic disorders, stroke, chronic migraine/others, depression, anxiety disorders, bipolar disorder and schizophrenia/others. Number of days (extension) in self-isolation was registered with a single question.

### Statistical analyses

Data were analyzed using mean and standard deviation (SD) or 95% confidence interval (95%CI) for continuous data and the raw numbers and % for categorical variables. Normality was checked with the Kolmogorov-Smirnov test. Due to the non-normal distribution, the mean changes (pre-versus during the pandemic) of MVPA levels and SB were evaluated using two generalized estimating equations models (GEE), one with changes in MVPA and one with change in SB as the dependent variable. The models were run testing the time effects (pre/during) and the interactions between time and the factors included in the model. The factors included in the models were (sex [male versus female], age [young adults {18-34 years} versus middle-age adults {35-54 years} versus older adults {55-65 years}], ethnicity [Caucasian versus Asian/Black/mixed/others], marital status [married versus single/divorced/widowed], employment [employed/students/military versus unemployed/retired], monthly household income [<R$2,005 versus R$2,005-R$8,640 versus R$8,641-R$11,261 versus >R$11,261], current smoking [yes versus no], alcohol consumption [yes versus no], self-reported previous diagnosis of any chronic diseases [yes versus no] or any mental disorders [yes versus no]). When the interaction between time and any factor was significant, the Bonferroni test was applied. The results of the models are presented using estimated marginal means and 95%CI. We also calculated the delta% change (pre to during), together with 95% CI as the effect size measure. The associations between the time (in days) in self-isolation and the change in MVPA and SB were tested using linear regression models. Days in self-isolation were collected as a continuous variable, and linear regression models were used to test the association of time in self-isolation with changes in MVPA and SB. The level of statistical significance was set at p-value < 0.05. All analyses were performed using SPSS (v. 21).

## Results

A total of 877 participants participated. The sample was predominantly comprised of women (n=635, 72.7%), young adults ranging from 18-34 years (n=471, 53.7%), Caucasians (n=669, 76.3%), singles (n=442, 50.9%), currently employed/students/military (n=723, 92.6%), with a monthly income ranging from R$2005 to R$8640 (n=364, 41.5%), non-smokers (n=836, 95.3%), currently consuming alcohol (n=605, 69.1%), with a self-reported previous diagnosis of a physical chronic disease (n=824, 94%) and without a self-reported previous diagnosis of a mental disorder (n=523, 59.6%). Participants were on average of 27.13 (6.57) days in self-isolation. The full details of the sample can be seen at Table 1.

**Table 1.** Sample characteristics

|  | Category/Mean<br>(Standard deviation) | Overall N=877* (%) |
| --- | --- | --- |
| **Sex** | Male | 238 (27.3) |
|  | Female | 635 (72.7) |
| **Age** | 18-24 years | 131 (14.9) |
|  | 25-34 years | 340 (38.8) |
|  | 35-44 years | 220 (25.1) |
|  | 45-54 years | 108 (12.3) |
|  | 55-64 years | 78 (8.9) |
| **Ethnicity** | Asian | 3 (0.3) |
|  | Black | 24 (2.7) |
|  | Mixed | 163 (18.6) |
|  | Caucasian | 669 (76.3) |
|  | Others | 16 (1.8) |
| **Marital Status** | Married | 363 (41.4) |
|  | Widowed | 4 (0.5) |
|  | Divorced | 60 (6.9) |
|  | Single | 442 (50.9) |
| **Employment** | Employed | 558 (63.6) |
|  | Unemployed | 43 (1.6) |
|  | Student | 235 (26.8) |
|  | Military | 19 (2.2) |
|  | Retired | 22 (2.5) |
| **Monthly household income** | <R\$1254 | 30 (3.4) |
| | R\$1255-R\$2004 | 94 (10.7) |
| | R\$2005-R\$8640 | 364 (41.5) |
| | R\$8641-R\$11261 | 139 (15.8) |
| | >R\$11262 | 250 (28.5) |
| **Current smoking** | No | 836 (95.3) |
|  | Yes | 41 (4.7) |
| **Current alcohol consumption** | No | 271 (30.9) |
|  | Yes | 605 (69.1) |
| **Self-reported previous diagnoses of physical disease** | No | 53 (6) |
|  | Yes | 824 (94) |
| **Self-reported previous diagnoses of mental disorder** | No | 523 (59.60) |
|  | Yes | 354 (40.4) |
| **Days in self isolation** | Mean (standard deviation) | 27.07 (6.71) |

\* Total sample with available data. Number of cases can be different for each variable due to missing cases (minimum=869)

### Mean changes in MVPA and SB (pre versus during pandemic)

A total of 432 (49.3%) of participants persisted to be active during the pandemic, 32 (3.6%) increased MVPA levels during the pandemic, 306 (34.9%) reduced MVPA levels during the pandemic, and 107 (12.2%) persisted to be inactive. We found an overall reduction of 64.28 (95% CI: 36.06 to 83.33) minutes per day on time spent in MVPA, corresponding to a reduction of 59.7% (95%CI: 35.64 to 82.21, p<0.001). The average time spent in SB increased 42.0% (95% CI: 31.74 to 52.50, p<0.001) during the pandemic, corresponding to additional 152.3 (95% CI: 111.9 to 192.7) minutes per day in SB. The mean time spent in MVPA and SB at baseline and during the pandemic can be found at figure 1.

**Figure 1.**
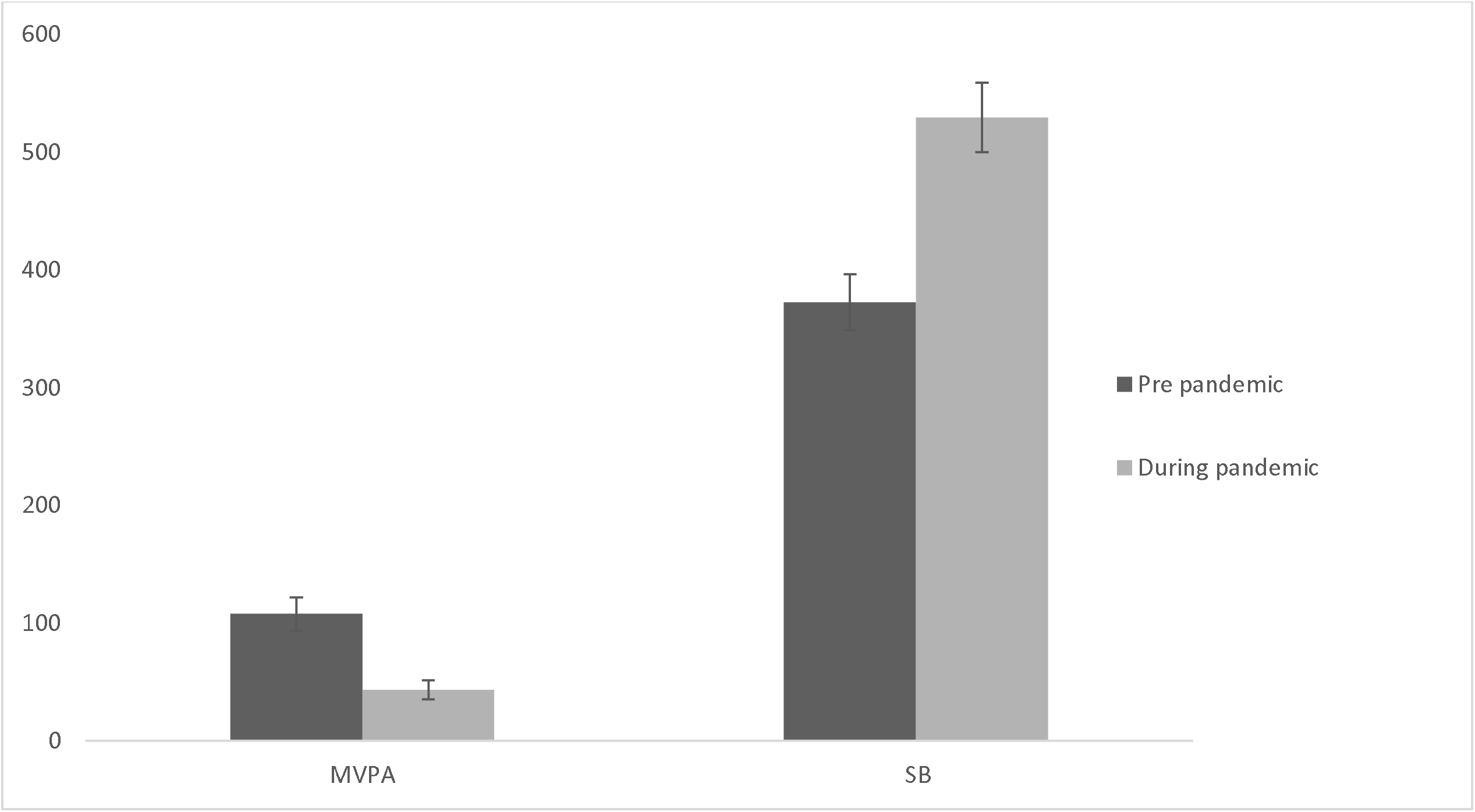
Moderate to vigorous physical activity and sedentary behavior, pre and during COVID-19 pandemic in 2020 in Brazil. Values are shown as the estimate marginal means, in minutes per day, of moderate to vigorous physical activity (MVPA) and sedentary behavior (SB) together with their standard error. Significant changes across time were found for MVPA (p<0.001) and SB (p<0.001).

### Correlates of MVPA change

Significant interactions in MVPA change were found for age (p=0.013), marital status (p=0.006) and employment (p=0.008). Bonferroni post hoc test found that young adults (mean change=-71.37, 95%CI: -99.76.51 to -42.98) and middle age adults (mean change=-66.76, 95%CI: -94.50 to -39.01) had greater decreases in time spent in MVPA when compared to older adults (mean change=-54.70, 95%CI: -86.25 to -23.16). Also, those not married (single/divorced/widowed) had greater reductions (mean change=- 75.50, 95%CI:-102.00 to -49.00) when compared to those married (mean change=- 53.05, 95%CI: -79.36 to -26.75), and those with an occupation (employed/student/military) had greater reductions (mean change=-78.69, 95%CI: - 105.21 to -52.16) when compared to those without occupation (unemployed/retired. Mean change=-49.87, 95%CI: -78.83 to -20.90). The detailed results of the MVPA model with mean changes can be seen in Table 2. Those not married (mean difference=22.08, 95%CI: 6.79 to 37.36, p=0.005), and with no occupation (mean difference=38.55, 95%CI: 14.08 to 63.03, p=0.002) had greater MVPA levels at baseline (pre and during pandemic means are shown in supplementary material 1). The number of days in self-isolation was not associated with changes in MVPA (unstandardized beta coefficient=0.234, 95%CI: −0.816 to 1.284, p=0.662, R^2^=0.00).

**Table 2.** Moderate to vigorous physical activity change (pre-post pandemic) in self-isolated adults during the SARS-CoV-2 pandemic in 2020 in Brazil

| Characteristics | Category | MVPA change (pre versus during) |  |  |  |  |  | Interaction<br>p value |
| --- | --- | --- | --- | --- | --- | --- | --- | --- |
|  |  | Mean<br>change in<br>minutes | 95% CI |  | Delta% | 95% CI |  |  |
| Sex | Male | -65.86 | -92.26 | -39.52 | -61.24% | -85.79% | -36.75% | 0.778 |
|  | Female | -62.71 | -89.37 | -36.05 | -58.18% | -82.91% | -33.44% |  |
| Age | Young adults (18-35 years) | **-71.37a** | **-99.76** | **-42.98** | **-72.68%** | **-101.51%** | **-43.77%** | **0.013** |
|  | Middle age adults (36-55 years) | **-66.76a** | **-94.50** | **-39.01** | **-64.53%** | **-91.34%** | **-37.07%** |  |
|  | Older adults (55-64 years) | **-54.70b** | **-86.25** | **-23.16** | **-45.08%** | **-71.08%** | **-19.08%** |  |
| Ethnicity | Black/Asian/Mixed/Others | -66.00 | -95.52 | -36.48 | -62.67% | -90.70% | -34.64% | 0.216 |
|  | White | -62.55 | -86.72 | -38.38 | -56.59% | -78.87% | -34.98% |  |
| Marital status | Single/divorced/widowed | **-75.50a** | **-102.00** | **-49.00** | **-63.60%** | **-85.93%** | **-41.28%** | **0.006** |
|  | Married | **-53.05b** | **-79.36** | **-26.75** | **-54.91%** | **-82.14%** | **-27.68%** |  |
| Employment | Employed/students/military | **-78.69a** | **-105.21** | **-52.16** | **-61.99%** | **-82.88%** | **-41.09%** | **0.008** |
|  | Unemployed/retired | **-49.87b** | **-78.83** | **-20.90** | **-56.42%** | **-89.19%** | **-23.64%** |  |
| Monthly household<br>income | <R\$2005 | -82.58 | -114.49 | -50.66 | -63.52% | -88.15% | -39.00% | 0.647 |
| | R\$2005-R\$8640 | -57.53 | -82.31 | -32.75 | -56.57% | -80.94% | -32.20% | |
| | R\$8641-R\$11261 | -56.72 | -89.26 | -24.19 | -58.06% | -91.38% | -24.76% | |
| | >R\$11261 | -60.28 | -88.92 | -31.63 | -59.44% | -87.69% | -31.19% | |
| Current smoking | Yes | -70.76 | -112.95 | -28.57 | -67.02% | -106.9% | -27.06% | 0.090 |
|  | No | -57.80 | -77.29 | -38.30 | -52.77% | -70.43% | -34.90% |  |
| Current alcohol<br>consumption | Yes | -58.39 | -84.85 | -31.93 | -57.09% | -82.97% | -31.22% | 0.378 |
|  | No | -70.16 | -97.22 | -43.10 | -62.06% | -85.29% | -38.12% |  |
| Self-reported previous<br>diagnosis of mental<br>disorders | Yes | -67.46 | -95.88 | -39.05 | -63.37% | -90.07% | -36.68% | 0.112 |
|  | No | -61.09 | -85.60 | -36.58 | -56.11% | -78.63% | -33.60% |  |
| Self-reported previous<br>diagnosis of physical<br>diseases | Yes | -67.42 | -89.62 | -45.23 | -59.64% | -79.28% | -40.01% | 0.803 |
|  | No | -61.13 | -96.90 | -25.63 | -59.77% | -94.74% | -25.63% |  |

Different letters mean significant differences according the Bonferroni post-hoc test (p<0.05).

**Table 3.**
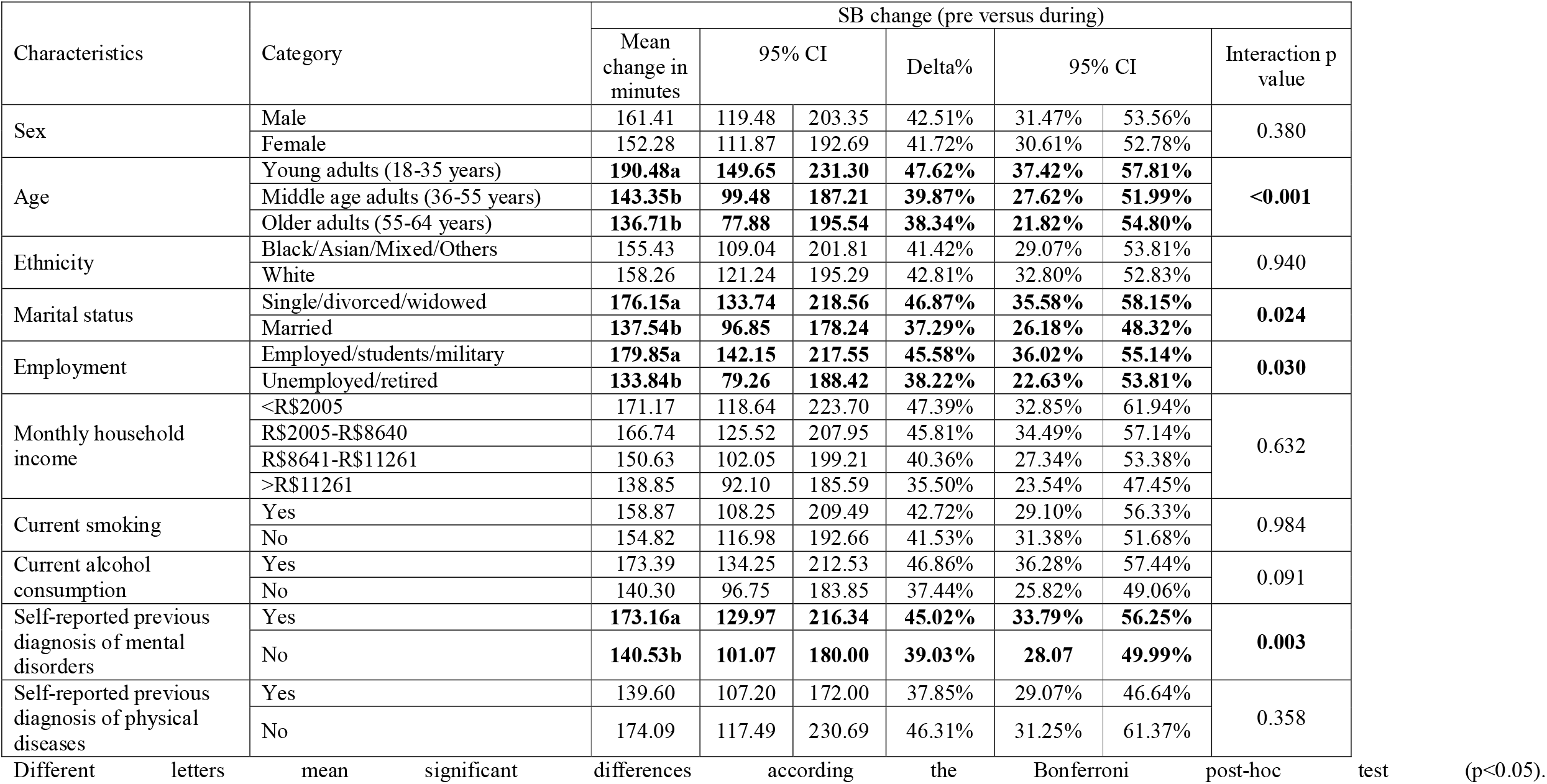
Sedentary behavior change (pre-post pandemic) in self-isolated adults during the SARS-CoV-2 pandemic in 2020 in Brazil

### Correlates of SB changes

The interactions found for the SB model were age (p<0.001), marital status (p=0.024), employment (p=0.03), and self-reported previous diagnosis of mental disorders (p=0.003). Young adults had greater increases (mean change=190.48, 95%CI: 149.65 to 231.30) in time spent in SB when compared to middle age (mean change=143.35 95%CI: 99.48 to 187.21) or older adults (mean change=136.71, 95%CI: 77.88 to 195.54). Also, greater increases in time spent in SB were found in those not married (mean change=176.15, 95%CI: 133.74 to 218.56) compared to those married (mean change=137.96, 95%CI: 96.85 to 178.24), in those with an occupation (mean change=179.85, 95%CI: 142.15 to 217.55) compared to those with no occupation (mean change=133.84, 95%CI: 79.26 to 188.42), and in those with a self-reported previous diagnosis of mental disorders (mean change=173.16, 95%CI: 129.97 to 216.34) compared to those with no history of mental disorders (mean change=140.53, 95%CI: 101.07 to 180.00). The detailed results of the SB model with interactions can be seen in Table 2. Younger adults spent more time in SB than middle-age adults (mean difference=39.99, 95%CI: 5.27 to 74.71, p=0.017), but not more than older adults (mean difference=43.28, 95%CI: -13.15 to 99.75, p=0.20). Those with a self-reported previous diagnosis of a mental disorder spent more time in SB at baseline (mean difference=24.09, 95%CI: 0.41 to 48.66, p=0.046) than those without (pre and during pandemic means are shown in supplementary material 2). The number of days in self-isolation was not associated with changes in SB (unstandardized beta coefficient=0.306, 95%CI: -1.732 to 2.345, p=0.77, R^2^=0.00).

## Discussion

The present study is, to the best of our knowledge, the first study demonstrating the impact of the self-isolation during the COVID-19 pandemic on Brazilians for self-reported MVPA and SB. Approximately 35.0% of the sample became insufficiently active during the self-isolation period. Only 3.6% became active with self-isolation. On average, there was a reduction of about 1 hour/day of time spent in MVPA, which corresponds to a reduction of 60.0% of their MVPA pre-pandemic levels. Participants reported spending 2.5 hours/day more in SB during the pandemic than before the pandemic, corresponding to an increase of 40.0%.

The reduction in MVPA levels in Brazilians is consistent with the findings of previous studies in other countries. For example, we found that roughly 35.0% of respondents became inactive during the self-isolation period as did about 50.0% in similar studies collected in France^11^, the USA^16^ and Australia^8^. Our results included reductions of 60.0% of the time spent in MVPA in Brazil, which is comparable to the reductions found in the USA, where there was a decrease of 47.0% on time spent in moderate PA^16^. Additionally, we observed an increase of about to 2.5 hours/day on time spent in SB, which is consistent with other studies that have found an increase of about 2 to 3 hours/day of SB in multiple countries^13, 21^. These findings highlight the urgent need for public health strategies to mitigate the impact of self-isolation on MVPA and SB.

Greater reductions in MVPA and increases in SB were found among younger adults, which is line with the findings from Italy and the UK^12, 14^. It is possible that this age group had fewer resources and greater difficulty coping with emotional responses to this situation^22^. Also, those not married and currently working had higher MVPA levels at baseline, but decreased their MVPA to similar levels to those not married and with no work during the pandemic. Of note, those currently employed might have reduced their commuting-related PA and have likely increased their SB time due to online meetings and activities. Lastly, those with a self-reported previous pandemic diagnosis of a mental disorder spent more time in SB and reported the greatest increases in time spent in SB during the pandemic. This finding is in accordance with a study in the UK^14^ that found a greater reduction in MVPA in people with depression. This finding is also consistent with previous studies showing that people with mental disorders have higher SB levels than people without mental disorders^23-25^, and suggests that self-isolation during the pandemic might be specifically detrimental to people with a previous diagnosis of a mental disorder.

There is ample evidence to justify making PA promotion a global public health priority during the coronavirus pandemic^3^. The COVID-19 pandemic appears to have impacted mental health globally, increasing rates of depression and anxiety symptoms and disorders^5^. On the other hand, those more active are less likely to develop mental disorders ^26-28^ and reported fewer symptoms in comparison to those with mental disorders ^29-32^. During the pandemic, cross-sectional evidence suggests that those with higher PA or lower SB levels are less likely to present depressive symptoms^33^. Promoting MVPA and reducing SB during the pandemic is also essential for physical health. Higher mortality due to COVID-19 is seen in those with clinical comorbidities such as hypertension, diabetes, and coronary heart disease^34^. Increasing time spent in MVPA and reducing time spent in SB seems to reduce the risk of development of multiple chronic diseases, including those associated with a higher risk of COVID-19 mortality^35^. Lastly, research is beginning to show that physical inactivity may be a risk factor for hospitalization due to COVID-19^36^, further underlining the potential importance of promoting PA during the pandemic.

The strengths of the manuscript are the large sample size of self-isolating Brazilians and the possibility to explore a variety of moderators. The present study has some limitations. First, MVPA and SB were assessed using self-reported questionnaires. Self-reported questionnaires are commonly associated with overestimations of MVPA^37^ and underestimation of SB^38^. Second, pre-pandemic MVPA and SB were assessed retrospectively, and both can be susceptible to memory bias. Third, the representativeness of the sample is limited. However, participants were drawn from 24 of the 27 federative units of Brazil, with more than half of participants from the Rio Grande do Sul, 14% from the Rio de Janeiro, and about 6% from Ceará. Also, some groups such as adults aging 55-64, Asian and Black people, and those with a household income lower than <R$1,254 are poorly represented. Fourth, we could not explore changes in light PA, such as walking. Also, we could not explore the changes in time spent on MVPA across the different PA domains (work/occupational, leisure, transportation, household). It is possible, for example, that some participants reduced the time spent in leisure, transportation, or work/occupational activities, but increased the time spent in household activities. This is important since we know that some mental health benefits are more likely to be associated with time spent in leisure activities^39^.

## Conclusion

Self-isolation during the pandemic significantly reduced time spent in MVPA and increased time spent in SB in Brazilian adults, particularly in younger adults, those who were single, and those who were employed. These findings highlight the urgent need of the adoption of public health strategies to address the impact of self-isolation during the COVID-19 pandemic on MVPA and SB.

## Data Availability

Data is available upon request.

## Acknowledgements

This study was part financed in part by the Coordenação de Aperfeiçoamento de Pessoal de Nível Superior - Brasil (CAPES) - Finance Code 001. André Werneck is supported by a São Paulo Research Foundation PhD scholarship (FAPESP process: 2019/24124-7). Joseph Firth is supported by a University of Manchester Presidential Fellowship (P123958) and a UK Research and Innovation Future Leaders Fellowship (MR/T021780/1). Brendon Stubbs is supported by a Clinical Lectureship (ICA-CL-2017-03-001) jointly funded by Health Education England (HEE) and the National Institute for Health Research (NIHR). Brendon Stubbs is part funded by the NIHR Biomedical Research Centre at South London and Maudsley NHS Foundation Trust. The views expressed are those of the author(s) and not necessarily those of the (partner organisation), the NHS, the NIHR or the Department of Health and Social Care. Mark Tully is partly supported by funding as Director of the Northern Ireland Public Health Research Network by the Research and Development Division of the Public Health Agency (Northern Ireland).

## References

1. World Health Organization. Coronavirus disease 2019 (COVID-19) : situation report, 179. 2020.

2. Ministério da Saúde. Boletim epidemiológico 05 - 20/03/2020. In: Saúde Md, ed. http://maismedicos.gov.br/images/PDF/2020_03_13_Boletim-Epidemiologico-05.pdf; 2020.

3. Sallis J, Adlakha D, Oyeyemi A, Salvo D. An international physical activity and public health research agenda to inform COVID-19 policies and practices. Journal of Sport and Health Science. 05/01 2020.

4. Ornell F, Schuch JB, Sordi AO, Kessler FHP. Pandemic fear and COVID-19: mental health burden and strategies. Brazilian Journal of Psychiatry. 2020.

5. Brooks SK, Webster RK, Smith LE, et al. The psychological impact of quarantine and how to reduce it: rapid review of the evidence. The Lancet. 2020.

6. Caspersen CJ, Powell KE, Christenson GM. Physical activity, exercise, and physical fitness: definitions and distinctions for health-related research. Public Health Rep. Mar-Apr 1985;100(2):126–131.

7. Gallo LA, Gallo TF, Young SL, Moritz KM, Akison LK. The impact of isolation measures due to COVID-19 on energy intake and physical activity levels in Australian university students. medRxiv. 2020:2020.2005.2010.20076414.

8. Stanton R, To Q, Khalesi S, et al. Depression, Anxiety and Stress during COVID-19: Associations with Changes in Physical Activity, Sleep, Tobacco and Alcohol Use in Australian Adults. Int J Environ Res Public Health. 06/07 2020;17:4065.

9. Lesser I, Nienhuis C. The Impact of COVID-19 on Physical Activity Behavior and Well-Being of Canadians. Int J Environ Res Public Health. 05/31 2020;17:3899.

10. Sekulic D, Blazevic M, Gilic B, Kvesic I, Zenic N. Prospective Analysis of Levels and Correlates of Physical Activity During COVID-19 Pandemic and Imposed Rules of Social Distancing; Gender Specific Study Among Adolescents from Southern Croatia. Sustainability. 2020;12(10):4072.

11. Deschasaux-Tanguy M, Druesne-Pecollo N, Esseddik Y, et al. Diet and physical activity during the COVID-19 lockdown period (March-May 2020): results from the French NutriNet-Sante cohort study. medRxiv. 2020:2020.2006.2004.20121855.

12. Giustino V, Parroco A, Gennaro A, Musumeci G, Palma A, Battaglia G. Physical Activity Levels and Related Energy Expenditure during COVID-19 Quarantine among the Sicilian Active Population: A Cross-Sectional Online Survey Study. Sustainability. 05/26 2020;12:4356.

13. Castañeda A, Arbillaga-Etxarri A, Gutiérrez-Santamaria B, Coca A. Impact of COVID-19 confinement on the time and intensity of physical activity in the Spanish population 2020.

14. Rogers N, Waterlow N, Brindle H, et al. Behavioural change towards reduced intensity physical activity is disproportionately prevalent among adults with serious health issues or self-perception of high risk during the UK COVID-19 lockdown 2020.

15. Lee Smith JM, Guillermo Felipe López-Sanchéz, Felipe Schuch, Igor Grabovac, Nicola Veronese, Ahmad Abufaraj, Cristina Coperchiona, Mark Tully. Prevalence and Correlates of Physical activity in a sample of UK adults observing social distancing during the COVID-19 pandemic. BMJ Open Sport & Exercise Medicine. 2020;in press.

16. Dunton G, Wang S, Do B, Courtney J. Early Effects of the COVID-19 Pandemic on Physical Activity in US Adults. 2020.

17. Dunton G, Do B, Wang S. Early Effects of the COVID-19 Pandemic on Physical Activity and Sedentary Behavior in US Children. 2020.

18. Meyer J, McDowell C, Lansing J, et al. Changes in physical activity and sedentary behaviour due to the COVID-19 outbreak and associations with mental health in 3,052 US adults 2020.

19. Tremblay MS, Aubert S, Barnes JD, et al. Sedentary behavior research network (SBRN)– terminology consensus project process and outcome. International Journal of Behavioral Nutrition and Physical Activity. 2017;14(1):75.

20. Cheval B, Sivaramakrishnan H, Silvio M, et al. Relationships between changes in selfreported physical activity, sedentary behaviours and health during the coronavirus (COVID-19) pandemic in France and Switzerland 2020.

21. Ammar A, Brach M, Trabelsi K, et al. Effects of COVID-19 Home Confinement on Eating Behaviour and Physical Activity: Results of the ECLB-COVID19 International Online Survey. Nutrients. 05/28 2020;12.

22. Scott SB, Sliwinski MJ, Blanchard-Fields F. Age differences in emotional responses to daily stress: the role of timing, severity, and global perceived stress. Psychol Aging. Dec 2013;28(4):1076–1087.

23. Schuch F, Vancampfort D, Firth J, et al. Physical activity and sedentary behavior in people with major depressive disorder: a systematic review and meta-analysis. Journal of affective disorders. 2017;210:139–150.

24. Vancampfort D, Firth J, Schuch F, et al. Physical activity and sedentary behavior in people with bipolar disorder: A systematic review and meta-analysis. J Affect Disord. Sep 1 2016;201:145–152.

25. Vancampfort D, Firth J, Schuch FB, et al. Sedentary behavior and physical activity levels in people with schizophrenia, bipolar disorder and major depressive disorder: a global systematic review and meta-analysis. World Psychiatry. Oct 2017;16(3):308–315.

26. Brokmeier LL, Firth J, Vancampfort D, et al. Does physical activity reduce the risk of psychosis? A systematic review and meta-analysis of prospective studies. Psychiatry Res. Nov 14 2019:112675.

27. Schuch FB, Vancampfort D, Firth J, et al. Physical Activity and Incident Depression: A Meta-Analysis of Prospective Cohort Studies. Am J Psychiatry. Jul 1 2018;175(7):631–648.

28. Schuch FB, Stubbs B, Meyer J, et al. Physical activity protects from incident anxiety: A meta-analysis of prospective cohort studies. Depression and anxiety. Sep 2019;36(9):846–858.

29. Ashdown-Franks G, Firth J, Carney R, et al. Exercise as medicine for mental and substance use disorders: A meta-review of the benefits for neuropsychiatric and cognitive outcomes. Sports Medicine. 2020;50(1):151–170.

30. Schuch FB, Vancampfort D, Richards J, Rosenbaum S, Ward PB, Stubbs B. Exercise as a treatment for depression: a meta-analysis adjusting for publication bias. Journal of psychiatric research. 2016;77:42–51.

31. Stubbs B, Vancampfort D, Hallgren M, et al. EPA guidance on physical activity as a treatment for severe mental illness: a meta-review of the evidence and Position Statement from the European Psychiatric Association (EPA), supported by the International Organization of Physical Therapists in Mental Health (IOPTMH). European psychiatry : the journal of the Association of European Psychiatrists. Oct 2018;54:124–144.

32. Stubbs B, Vancampfort D, Rosenbaum S, et al. An examination of the anxiolytic effects of exercise for people with anxiety and stress-related disorders: A meta-analysis. Psychiatry Res. Mar 2017;249:102–108.

33. Schuch FB, Bulzing RA, Meyer J, Vancampfort D, Firth J. Associations of moderate to vigorous physical activity and sedentary behavior with depressive and anxiety symptoms in self-isolating people during the COVID-19 pandemic: A cross-sectional survey in Brazil.

34. Zhou F, Yu T, Du R, et al. Clinical course and risk factors for mortality of adult inpatients with COVID-19 in Wuhan, China: a retrospective cohort study. The Lancet. 2020/03/28/ 2020;395(10229):1054–1062.

35. Pedersen BK, Saltin B. Exercise as medicine–evidence for prescribing exercise as therapy in 26 different chronic diseases. Scandinavian journal of medicine & science in sports. 2015;25:1–72.

36. Hamer M, Kivimäki M, Gale CR, Batty GD. Lifestyle risk factors, inflammatory mechanisms, and COVID-19 hospitalization: A community-based cohort study of 387,109 adults in UK. Brain Behav Immun. Jul 2020;87:184–187.

37. Lee PH, Macfarlane DJ, Lam TH, Stewart SM. Validity of the international physical activity questionnaire short form (IPAQ-SF): A systematic review. International Journal of Behavioral Nutrition and Physical Activity. 2011/10/21 2011;8(1):115.

38. Prince S, Cardilli L, Reed J, et al. A comparison of self-reported and device measured sedentary behaviour in adults: a systematic review and meta-analysis. International Journal of Behavioral Nutrition and Physical Activity. 03/04 2020;17:1–17.

39. Teychenne M, White RL, Richards J, Schuch FB, Rosenbaum S, Bennie JA. Do we need physical activity guidelines for mental health: What does the evidence tell us? Mental Health and Physical Activity. 2020;18:100315.

